# Multimodal MRI Characterization of Nucleus Basalis of Meynert Degeneration: Structural Atrophy and Free-water Diffusion in Parkinson’s Disease Cognitive Impairment

**DOI:** 10.64898/2026.06.08.26355183

**Authors:** Ahmed Negida, Ananna Zaman, Kathryn A. Wyman-Chick, Razan Hallak, Cameron Miller-Patterson, Brian D. Berman, Edward Ofori, Matthew J. Barrett

**Affiliations:** Parkinson and Movement Disorder Center, Department of Neurology, Virginia Commonwealth University, Richmond, VA, United States of America; Department of Neurology, HealthPartners, Golden Valley, Minnesota, USA; College of Health Solutions, Arizona State University, Phoenix, AZ, USA

**Keywords:** Parkinson’s disease, cholinergic basal forebrain, nucleus basalis of Meynert, free water, cognitive impairment, diffusion MRI

## Abstract

**Background:** Cognitive impairment in Parkinson’s disease (PD) is linked to degeneration of the cholinergic basal forebrain, particularly cholinergic nucleus 4 (Ch4) in the nucleus basalis of Meynert. Structural and diffusion MRI separately detect this degeneration, but few studies have combined these modalities across the PD cognitive spectrum.

**Methods:** We analyzed 92 participants: 14 healthy controls (HC), 35 PD with normal cognition (PD-NC), 33 with mild cognitive impairment (PD-MCI), and 10 with dementia (PDD). For Ch4 and cholinergic nuclei 1, 2, and 3 (Ch1-3) in the medial septal/diagonal band complex, we determined TIV-normalized gray matter density (GMD) and free-water (FW) fraction. We evaluated group differences, cognitive correlations, adjusted multivariable regression, and exploratory ROC discrimination.

**Results:** Ch4 GMD was significantly lower in PDD compared to PD-MCI (p=0.007), PD-NC (p<0.001), and HC (p<0.001). Ch4 GMD was also lower in PD-MCI versus HC (p=0.028); the PD-MCI versus PD-NC difference was not significant after correction (p=0.074). Ch1-3 GMD was lower in PDD versus PD-NC (p=0.008) and HC (p=0.009). Ch4 and Ch1-3 FW were elevated in PDD versus all other groups (all p<0.01). Among PD patients (n=78), MoCA was positively correlated with Ch4 GMD (ρ=0.49) and Ch1-3 GMD (ρ=0.42) and negatively correlated with Ch4 FW (ρ=−0.51) and Ch1-3 FW (ρ=−0.40; all p<0.001). In the full four-metric model, Ch4 GMD and Ch4 FW were the only independent basal forebrain predictors (Ch4 GMD β=+2.04, p<0.001; Ch4 FW β=−1.46, p=0.005) of MoCA score. The combined Ch4 GMD + Ch4 FW model showed high discrimination for PDD versus non-demented PD (AUC=0.934; optimism-corrected AUC=0.925).

**Conclusions:** Structural and free-water diffusion MRI provide complementary information about Ch4 degeneration in PD. The combined Ch4 model showed promising exploratory discrimination of PDD; validation in larger independent samples is needed.

## Introduction

Cognitive impairment is one of the most prevalent and debilitating nonmotor symptoms of Parkinson’s disease (PD). Mild cognitive impairment in PD (PD-MCI), which is present in approximately 25–30% of patients (Litvan et al., 2011), often develops into PD dementia (PDD), with up to 75% of patients experiencing dementia at 20 years (Gallagher et al., 2024). In addition to being common, cognitive impairment in PD is a major determinant of quality of life and caregiver burden (Aarsland et al., 2021).

Degeneration of the cholinergic basal forebrain, particularly the nucleus basalis of Meynert, is a major contributor to cognitive decline in PD (Bohnen & Albin, 2011). Cholinergic nucleus 4 (Ch4), the cholinergic constituent of the nucleus basalis of Meynert, provides the primary cholinergic innervation to the neocortex and amygdala, and its degeneration has been consistently linked to cognitive impairment in PD (Barrett et al., 2019; Ray et al., 2018; Schulz et al., 2018). The medial septal nucleus (cholinergic nucleus 1; Ch1) and vertical limb of the diagonal band of Broca (cholinergic nucleus 2; Ch2) provide cholinergic projections to the hippocampus and have been implicated in memory impairment in PD (Gargouri et al., 2019). The horizontal limb of the diagonal band of Broca (cholinergic nucleus 3; Ch3) provides cholinergic innervation to the olfactory bulb and piriform cortex (Mesulam et al., 1983).

Most MRI studies evaluating the cholinergic basal forebrain in PD have focused on structural sequences to assess macrostructural atrophy using volumetric measures (Pasquini et al., 2021). While these studies have demonstrated associations between cholinergic basal forebrain volume loss and cognitive decline, macrostructural atrophy may only occur at a later stage of neurodegeneration. Diffusion MRI has the potential to identify microstructural changes that may precede or accompany atrophy during neurodegeneration. An earlier study using DTI found that Ch4 mean diffusivity (MD) was increased in PD with cognitive impairment compared to PD with normal cognition and higher Ch4 MD was an independent predictor of cognitive decline in PD while Ch4 gray matter volume was not (Schulz et al., 2018). Gargouri et al. reported that higher Ch3 and Ch4 DTI-derived MD, axial diffusivity (AD), and radial diffusivity (RD) correlated with worse executive function, while higher Ch1 and Ch2 MD and RD correlated with worse delayed recall (memory) and visuospatial performance on a copy task (Gargouri et al., 2019). Perhaps as a result of using a global region of interest for the cholinergic basal forebrain, a subsequent study found no relationship between MD of the cholinergic basal forebrain and cognitive measures (Grothe et al., 2021).

One of the challenges of conventional diffusion tensor imaging (DTI) metrics is confounding introduced by partial volume effects from cerebrospinal fluid contamination, which is particularly problematic in periventricular structures such as the cholinergic basal forebrain. The free water (FW) imaging framework, developed by Pasternak et al.(Pasternak et al., 2009), addresses this limitation by modeling each voxel as a combination of a tissue compartment and a free-water compartment. This bi-tensor model yields a FW fraction reflecting extracellular water content and tissue-corrected diffusion metrics that more accurately characterize parenchymal microstructure. Elevated FW has been interpreted as reflecting neuroinflammation, neurodegeneration, or atrophy-related changes (Ofori et al., 2015; Pasternak et al., 2009).

Thus, isolating the extracellular free water (FW) fraction using DTI may provide a more biologically meaningful measure of microstructural degeneration than conventional diffusivity measures such as MD, AD, and RD, and a few studies have applied these methods to the cholinergic basal forebrain in PD. Two prior studies reported increased Ch4 FW in PD with cognitive impairment compared to PD with normal cognition (Ray et al., 2023; Zhang et al., 2024). In one higher FW was correlated with worse global cognition (Ray et al., 2023), and in the other higher Ch4 FW was correlated with worse executive function, processing speed, working memory, and delayed recall. Demonstrating the value of measuring Ch4 FW over volume, a study in isolated REM sleep behavior disorder found that elevated Ch4 FW but not Ch4 volume was associated with a greater risk of conversion to dementia with Lewy bodies over PD (Haddad et al., 2025).

To date, there have been limited efforts to determine whether combining the imaging modalities of structural and diffusion MRI can yield a more biologically complete indicator of neurodegeneration in the cholinergic basal forebrain relevant to cognitive impairment in PD. The current study aimed to expand on prior work by investigating and integrating structural T1 MRI and FW of Ch4 and cholinergic nucleus 1, 2, and 3 (Ch1-3) in a cohort of PD participants with normal cognition (PD-NC), mild cognitive impairment (PD-MCI) and dementia (PDD). Our objectives were 1) to determine the pattern of differences in Ch4 and Ch1-3 structural and diffusion measures between PDD, PD-MCI, PD-NC, and healthy controls, 2) to determine the relationships between Ch4 and Ch1-3 structural and diffusion measures and cognitive measures in PD, and 3) to determine which combination of Ch4 and Ch1-3 MRI metrics were the most predictive of cognitive performance and were the most accurate for identifying cognitive status in PD.

## Methods

### Study population

This cross-sectional single-center study included 92 participants recruited at the Parkinson and Movement Disorder Center, Virginia Commonwealth University including 14 health controls and 78 people with PD. All PD participants met clinical diagnostic criteria for established or probable PD (Postuma et al., 2015). Core exclusion criteria were prior deep brain stimulation, any history of neurosurgical intervention, and the presence of other significant neurological disorders aside from PD. All study procedures were approved by the Virginia Commonwealth University Institutional Review Board, and all participants provided written informed consent (or consent was obtained from a legally authorized representative when applicable).

### Clinical and neuropsychological assessments

All participants completed the Movement Disorder Society–Unified Parkinson’s Disease Rating Scale (MDS-UPDRS). The Montreal Cognitive Assessment (MoCA) was administered to assess global cognition, and every participant also completed a comprehensive neuropsychological testing battery with two neuropsychological tests in each domain. To assess executive function, participants completed the Trail Making Test Part B (TMT-B) and FAS (controlled oral word association). Heaton norms, adjusted for age, sex, years of education, and race, were applied to generate T-scores for TMT-B and FAS (Heaton et al., 2004). To assess attention, the Symbol Digit Modalities Test (SDMT), Brief Test of Attention (BTA), fourth edition (WAIS-IV) Digit Span Forward (DSF), and Digit Vigilance Test (DVT) form 6 were administered. WAIS-IV DSF scaled scores were calculated using age-standardized norms (Wechsler, 2008). For the Symbol Digit Modalities Test (SDMT), norms adjusted for age and years of education were used to obtain z-scores (Smith, 1982). Heaton norms, adjusted for age, sex, years of education, and race, were also used to generate T-scores for Digit Vigilance Test (DVT) total time (Heaton et al., 2004).

Brief Test of Attention z-scores were derived using normative data standardized for age and education (Schretlen et al., 1996). To assess memory, participants completed the Hopkins Verbal Learning Test-Revised (HVLT-R) form 1 and the Brief Visuospatial Memory Test-Revised (BVMT-R) form 1. Age-adjusted normative data for the Hopkins Verbal Learning Test-Revised (HVLT-R) and the Brief Visuospatial Memory Test-Revised (BVMT-R) were applied to derive delayed recall T-scores for each test (Benedict, 1997; Benedict et al., 1998). To assess language, participants completed the Category fluency test (the Animal Naming Test) and the Boston Naming Test (BNT) 15-item short form. For the BNT 15-item short form, age-adjusted norms were used to compute individual z-scores (Mack et al., 1992). Heaton norms were used to generate a T-score for the Animal Naming Test (Heaton et al., 2004). To assess visuospatial function, the 15-Item Judgment of Line Orientation form H, BVMT-R Copy Trial, and WMS-IV VR figural Copy Trial were administered. For visuospatial function, a 30-item JLO equivalent raw score was estimated from the 15-item raw score using a linear conversion (raw score × 2). This converted raw score was then used to obtain an age-adjusted JLO scaled score (Woodard et al., 1998). JLO z-scores were derived from the scaled scores using a mean of 12.3 and a standard deviation of 2.3 (Woodard et al., 1998). The BVMT-R Copy Trial was administered, and raw scores were obtained. The BVMT-R copy trial was not administered to 12 participants overall, the MoCA copy score (cube copying task) was used to supplement any missing copy trial scores.

Due to the severity of their cognitive impairment, seven participants were unable to complete TMT–B; five could not complete the SDMT, and five were unable to complete the JLO. For those who could not complete TMT–B, the lowest possible T-score was assigned based on Heaton norms adjusted for age, sex, years of education, and race (Heaton et al., 2004). For participants unable to complete the SDMT, a raw score of zero was assigned and converted to a z-score using norms standardized for age and education. Similarly, for those who could not complete the JLO, a raw score of zero was assigned and converted to an age-adjusted scaled score using JLO norms (Woodard et al., 1998). All test scores were standardized and converted to z-scores, with the exception of the Copy Trial and MoCA copying task. Composite domain z-scores were derived by averaging the available z-scores within each domain, except for the visuospatial domain in which only the JLO z-score was available. A global cognitive composite z-score was calculated as the mean of the five-domain z-scores (memory, executive, language, attention, and visuospatial).

### Cognitive classification

Performance ≥1.5 standard deviations below the normative mean were considered impaired for all neuropsychological measures except for the BVMT-R copy trial. Scores less than 11 on BVMT-R Copy Trial and less than 38 on VR figural copy trial were considered impaired (Powell et al., 2022). For the 12 participants with a missing copy trial raw score, a MoCA copy score of 0 was considered impaired. PD participants were designated as having normal cognition (PD-NC) if they scored greater than 25 on the MoCA and were not impaired on more than 1 neuropsychological tests. Consistent with Movement Disorder Society Level II PD-MCI criteria, participants with PD who scored less than 26 on the MoCA or demonstrated impairment on two or more neuropsychological tests were classified as PD-MCI (Litvan et al., 2012). Participants were designated as having PDD if they met standard criteria (Emre et al., 2007). All PDD participants were also confirmed to have impairment on 2 or more neuropsychological tests.

### MRI acquisition

All participants underwent magnetic resonance imaging on the same 3T Philips scanner (Philips Healthcare, Best, The Netherlands). T1-weighted images were acquired using a three-dimensional turbo field echo (3D-TFE) sequence with the following parameters: repetition time (TR) = 6.66 ms, echo time (TE) = 3.0 ms, inversion time (TI) = 1060 ms, field of view (FOV) = 240 × 256 × 225 mm, acquisition matrix = 256 × 256, 225 sagittal slices, and isotropic voxel size of 1.0 × 1.0 × 1.0 mm. Diffusion-weighted images were acquired using a spin-echo echo-planar imaging sequence with multi-band acceleration factor 3 (MB3): echo time (TE) = 115 ms, field of view = 224 × 224 mm, acquisition matrix = 144 × 144, 60 axial slices, voxel size = 1.56 × 1.56 × 2.20 mm, and phase-encoding direction posterior-to-anterior. Diffusion gradients were applied along 128 non-collinear directions across two b-value shells: 64 directions at b = 1000 s/mm² and 64 directions at b = 3000 s/mm², plus one non-diffusion-weighted volume (b = 0 s/mm²), yielding 129 total volumes. Reverse phase-encoded b = 0 volumes were additionally acquired for susceptibility-induced distortion correction.

### Structural T1 MRI processing

T1-weighted images were processed using the Computational Anatomy Toolbox (CAT12) within SPM12. Prior to processing, the origin of each image was manually reoriented to the anterior commissure. Within CAT12, images were denoised using spatial-adaptive non-local means (SANLM) denoising and a Markov random field approach, bias corrected, spatially normalized to standard stereotactic space with an affine registration, and locally intensity-transformed.

Normalized images were segmented into gray matter, white matter, and cerebrospinal fluid using the Adaptive Maximum A Posteriori (AMAP) technique. Tissue priors were used for spatial normalization, skull-stripping, and initial segmentation estimation within the AMAP segmentation. Partial-volume estimation was performed to account for voxels containing more than one tissue type. The Diffeomorphic Anatomic Registration Through Exponentiated Lie (DARTEL) algorithm and Geodesic Shooting were used to register segmented images to standard Montreal Neurological Institute (MNI) space. Segmented images were modulated by the amount of volume change from spatial registration to preserve the total amount of gray matter.

Ch4 (nucleus basalis of Meynert, NBM) and Ch1-3 (medial septal nucleus and diagonal band of Broca) gray matter density values were extracted in MNI space from CAT12 modulated normalized gray matter maps using probability-weighted means within the Zaborszky et al. probabilistic basal forebrain atlas (Zaborszky et al., 2008). Values were normalized by total intracranial volume (TIV) to control for head size and multiplied by 1000 for interpretability; the resulting metric represents probability-weighted modulated gray matter volume per ROI scaled by TIV. Throughout this article, we refer to this TIV-normalized probability-weighted modulated gray matter measure as Ch4/Ch1-3 GMD.

### Diffusion Tensor Imaging processing

Diffusion-weighted volumes underwent MP-PCA denoising (Veraart et al., 2016), Gibbs ringing correction (Kellner et al., 2016), eddy current and motion correction (FSL eddy with topup, PE direction: PA) (Andersson & Sotiropoulos, 2016), and N4 bias field correction (Tustison et al., 2010). Brain masks were generated using dwi2mask (MRtrix3 3.0.4). Free-water (FW) fraction was estimated using the bi-tensor model with Beltrami-regularized gradient descent (Pasternak et al., 2009), implemented in the fwe package (Golub et al., 2021). For multi-shell acquisitions, the b = 0 and b = 1000 s/mm² shells were extracted to match the single-shell bi-tensor framework used in prior basal forebrain studies (Ray et al., 2023; Wu et al., 2024; Xu et al., 2025; Zhang et al., 2024). The model separates the diffusion signal into a tissue compartment (anisotropic tensor) and a free-water compartment (isotropic diffusion, D = 3.0 × 10LJ³ mm²/s), yielding voxelwise maps of FW fraction. Regularization parameters followed default settings (100 iterations, learning rate 0.0005, hybrid initialization).

T1-weighted images were reoriented to standard orientation (FSL fslreorient2std) and skull-stripped using ANTs template-based brain extraction (antsBrainExtraction.sh; ANTs 2.5.4) with the OASIS T_template0 and accompanying brain-cerebellum probability mask. Tissue segmentation was performed with FSL FAST. T1-to-MNI152 (2 mm) normalization was performed using ANTs symmetric diffeomorphic registration (antsRegistrationSyN.sh) with three stages: rigid (6-DOF), affine (12-DOF), and SyN diffeomorphic nonlinear registration with the cross-correlation metric (Avants et al., 2011). DWI-to-T1 alignment was achieved via boundary-based registration (FSL epi_reg). The resulting warp chain (DWI → T1 → MNI) was used to project all diffusion metric maps into MNI152 (2 mm) standard space. Cholinergic basal forebrain nuclei were defined using the Zaborszky cytoarchitectonic probabilistic atlas (Zaborszky et al., 2008), resampled to 2 mm MNI space.

Two regions of interest were examined: Ch4 (nucleus basalis of Meynert, NBM) and Ch1-3 (medial septum and diagonal band complex). Following Ray et al. (Ray et al., 2023), all diffusion metric maps were warped to MNI standard space (rather than warping atlas ROIs to native diffusion space) to avoid spatial distortions from head positioning. Within each ROI, FA conditioning was applied to exclude voxels with FA > 0.3 (white matter contamination) or FA < 0.05 (CSF contamination), retaining only voxels within the gray matter range characteristic of basal forebrain nuclei (Schulz et al., 2018). ROI probability-weighted means were computed using the atlas probability values as weights.

### Statistical analysis

Statistical analyses were performed using Python v3 and Stata MP v18. We used Kruskal-Wallis tests for omnibus four-group comparisons and Mann-Whitney U tests with Bonferroni correction for pairwise between-group comparisons of imaging metrics. Correlation between basal forebrain imaging metrics (Ch4 and Ch1-3 GMD and FW) and cognitive measures were assessed with Spearman rank correlations with Benjamini–Hochberg false-discovery-rate correction and graphically presented in a heatmap.

To clarify which basal forebrain measures were independent predictors of cognition, a series of multivariable linear regression models were fit for each cognitive outcome (MoCA total, global composite z-score, and each of five cognitive domain z-scores). Because the four basal forebrain metrics (Ch4 GMD, Ch1-3 GMD, Ch4 FW, Ch1-3 FW) were correlated (Pearson r ranging from −0.63 to +0.79; all variance inflation factors < 5) and entering all four simultaneously can produce coefficient instability and suppression effects, a tiered modeling strategy was used so that each model addressed a distinct question. All models were adjusted for age, sex, and education; basal forebrain predictors were z-standardized within the PD sample so that β represents the change in outcome per 1-SD increase in the imaging predictor. All within-PD analyses used the 78 PD participants.

The tiered modeling strategy proceeded as follows. For Tier 1 (univariate-adjusted), each basal forebrain metric was entered alone with covariates. For Tier 2a (within-Ch4) and Tier 2b (within-Ch1-3), GMD and FW were paired within the same nucleus to test whether microstructure adds to structure (Ch4 GMD with Ch4 FW; Ch1-3 GMD with Ch1-3 FW). For Tier 3a (between-nuclei GMD) and Tier 3b (between-nuclei FW), the two structural metrics were entered together (Ch4 GMD with Ch1-3 GMD) and the two microstructural metrics together (Ch4 FW with Ch1-3 FW), to test which nucleus dominates within each modality. For Tier 4 (full 4-metric), all four metrics were entered together as a full model, reported for completeness with the caveat that individual coefficients in this model are subject to collinearity-driven instability. ROC analyses were exploratory; AUC point estimates are reported with 95% bootstrap confidence intervals (2,000 resamples) and Harrell bootstrap optimism correction (1,000 resamples).

## Results

### Participant characteristics

The study population included 92 participants with complete multimodal basal forebrain imaging: 14 healthy controls (HC), 35 PD with normal cognition (PD-NC), 33 PD with mild cognitive impairment (PD-MCI), and 10 PD with dementia (PDD). Demographic and clinical characteristics are presented in Table 1. Groups did not differ significantly in age (p=0.105), sex (p=0.252), or years of education (p=0.962). MDS-UPDRS Part III motor scores and MDS-UPDRS total scores were significantly higher in PDD compared to PD-NC and PD-MCI (all p<0.005). As expected, MoCA scores and cognitive domain z-scores differed significantly across groups (all p≤0.003). MoCA was significantly different in all pairwise group comparisons (all p<0.001) except for PD-NC vs HC.

**Table 1.** Demographic and clinical characteristics of participants by cognitive group.

| Variable | HC (n=14) | PD-NC (n=35) | PD-MCI (n=33) | PDD (n=10) | p-value |
| --- | --- | --- | --- | --- | --- |
| Age, years | 70.0 (60.6–73.0) | 68.4 (65.7–71.0) | 69.3 (66.9–74.0) | 74.5 (69.0–75.8) | 0.105 |
| Education, years | 16.0 (15.2–18.0) | 17.0 (16.0–18.0) | 16.0 (15.0–18.0) | 18.0 (15.5–18.0) | 0.962 |
| Sex, male n (%) | 6 (42.9%) | 17 (48.6%) | 22 (66.7%) | 7 (70.0%) | 0.252 |
| MDS-UPDRS Part III | — | 24.0 (20.5–38.5) | 33.0 (23.0–40.0) | 46.5 (40.2–58.0) | 0.002 |
| MDS-UPDRS total | — | 45.0 (37.0–64.0) | 53.0 (41.0–74.0) | 94.0 (86.0–105.0) | <0.001 |
| MoCA total | 28.5 (27.0–29.0) | 28.0 (26.5–29.0) | 25.0 (23.0–26.0) | 18.5 (16.5–19.8) | <0.001 |
| Memory z | 0.6 (0.2 to 0.9) | 0.6 (0.0 to 0.9) | −1.0 (−1.6 to −0.1) | −2.5 (−3.0 to −2.3) | <0.001 |
| Executive z | 0.7 (0.2 to 1.2) | 0.1 (−0.2 to 0.7) | −0.6 (−1.4 to −0.2) | −2.7 (−3.0 to −2.3) | <0.001 |
| Language z | 0.7 (0.3 to 0.7) | 0.4 (0.2 to 0.8) | 0.0 (−0.8 to 0.3) | −1.6 (−2.0 to −1.2) | <0.001 |
| Attention z | 0.8 (0.3 to 0.9) | 0.0 (−0.4 to 0.6) | −0.4 (−0.9 to −0.1) | −1.6 (−1.9 to −0.9) | <0.001 |
| Visuospatial z* | 0.3 (−0.1 to 0.7) | 0.3 (−1.0 to 0.7) | −0.1 (−1.9 to 0.7) | −3.2 (−3.6 to −2.0) | 0.003 |
| Ch4 GMD (TIV-normalized) | 52.5 (49.8–55.1) | 52.1 (49.3–53.5) | 49.5 (47.0–50.7) | 43.8 (40.6–44.8) | <0.001 |
| **Ch1-3 GMD**<br>**(TIV-**<br>**normalized)** | 34.1 (33.3–<br>36.0) | 34.3 (32.7–36.4) | 32.7 (31.7–34.5) | 30.3 (29.2–32.6) | 0.001 |
| **Ch4 FW fraction** | 0.447 (0.425–<br>0.480) | 0.467 (0.443–<br>0.482) | 0.498 (0.473–<br>0.546) | 0.583 (0.547–0.601) | <0.001 |
| **Ch1-3 FW**<br>**fraction** | 0.449 (0.405–<br>0.478) | 0.454 (0.434–<br>0.473) | 0.492 (0.441–<br>0.511) | 0.591 (0.554–0.611) | <0.001 |

Data are presented as median (interquartile range) unless otherwise indicated. P-values were derived from Kruskal-Wallis tests for continuous variables and Fisher's exact test for the categorical variable (sex). \* Only 76 PD participants had visuospatial z-scores. Abbreviations: FW, free water fraction; GMD, gray matter density; HC, healthy controls; JLO, Judgment of Line Orientation; MDS-UPDRS, Movement Disorder Society–Unified Parkinson's Disease Rating Scale; MoCA, Montreal Cognitive Assessment; PD-MCI, Parkinson's disease with mild cognitive impairment; PD-NC, Parkinson's disease with normal cognition; PDD, Parkinson's disease with dementia; TIV, total intracranial volume.

### Cholinergic basal forebrain gray matter density and free water

Ch4 GMD differed significantly across the four groups (Kruskal-Wallis p < 0.001; Figure 1A). Bonferroni-corrected post-hoc Mann-Whitney U tests revealed that PDD had significantly lower Ch4 GMD than PD-MCI (p=0.007), PD-NC (p<0.001), and HC (p<0.001), and PD-MCI had significantly lower Ch4 GMD than HC (p=0.028); the PD-MCI vs PD-NC comparison did not reach Bonferroni-corrected significance (p=0.074). Ch1-3 GMD was also significantly different across groups (Kruskal-Wallis p < 0.001; Figure 1B). PDD had significantly lower Ch1-3 GMD than PD-NC (p=0.008) and HC (p=0.009); other pairwise comparisons did not reach Bonferroni-corrected significance.

**Figure 1.**
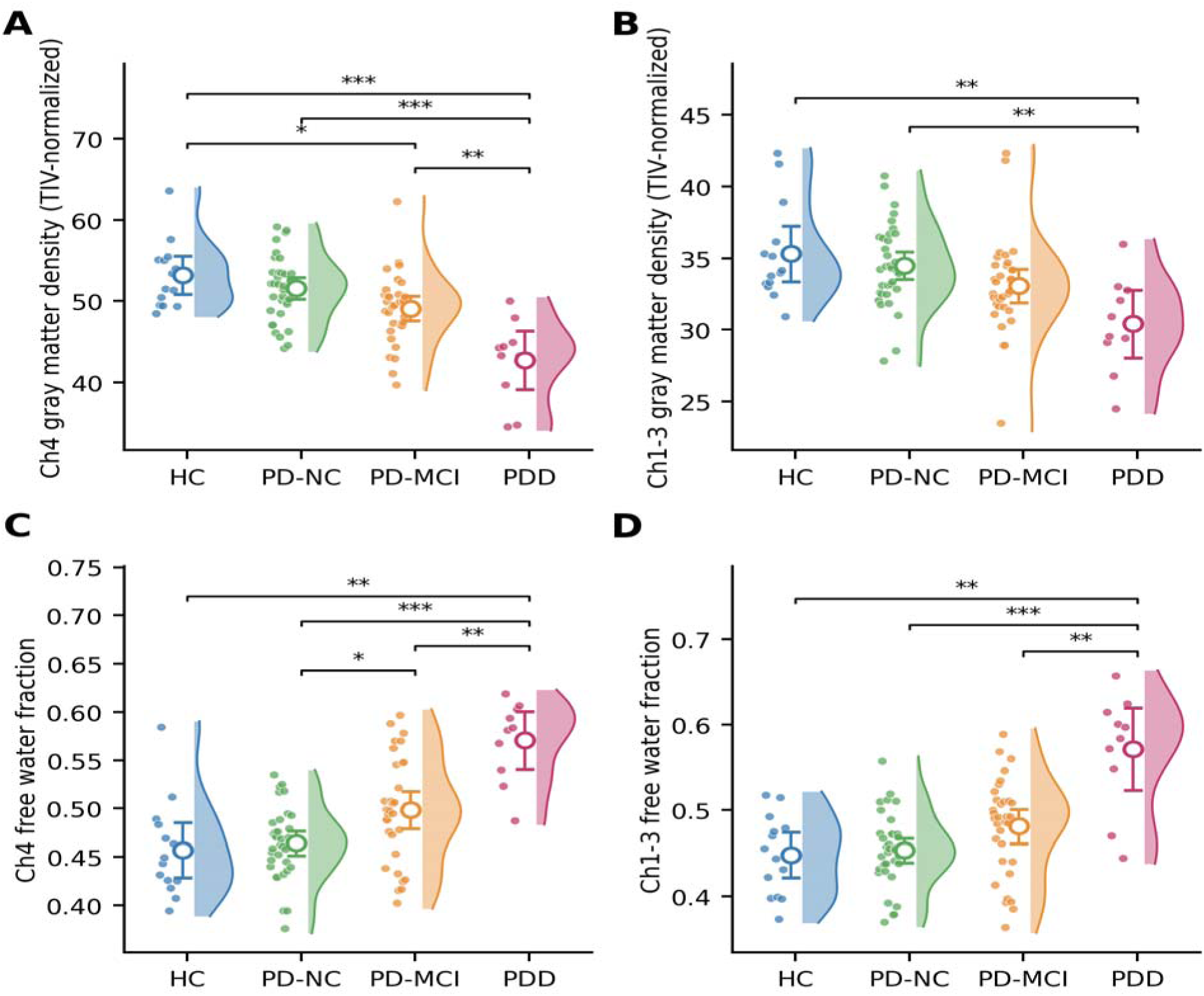
Basal forebrain gray matter density (GMD) and free water fraction (FW) across participant groups. (A) Ch4 GMD (TIV-normalized). (B) Ch1-3 GMD (TIV-normalized). (C) Ch4 FW. (D) Ch1-3 FW. Raincloud plots show the probability density (half-violin), individual data points (jittered strip), and median with interquartile range. Pairwise comparisons performed using Mann-Whitney U tests with Bonferroni correction for six comparisons across the four groups (HC, PD-NC, PD-MCI, PDD). *p<0.05, **p<0.01, ***p<0.001; only statistically significant pairwise comparisons are shown. HC, healthy controls; PD-NC, PD with normal cognition; PD-MCI, PD with mild cognitive impairment; PDD, PD with dementia.

Ch4 FW was significantly different between groups (p < 0.001; Figure 1C). PDD showed significantly higher Ch4 FW compared to PD-MCI (p = 0.006), PD-NC (p < 0.001), and HC (p = 0.002). A similar pattern was observed for Ch1-3 FW (p < 0.001; Figure 1D), with PDD showing significantly elevated values compared to PD-MCI (p = 0.007), PD-NC (p < 0.001), and HC (p = 0.005).

### Correlations with cognitive performance among PD participants

Among PD participants with complete multimodal imaging (N=78), MoCA total score was positively correlated with Ch4 GMD (ρ=0.49, p<0.001) and Ch1-3 GMD (ρ=0.42, p<0.001) and negatively correlated with Ch4 FW (ρ=−0.51, p<0.001) and Ch1-3 FW (ρ=−0.40, p<0.001) (Figure 2). Similarly, Ch4 GMD showed the strongest positive correlation with the global cognitive composite z-score (ρ=0.48, p<0.001), and Ch4 FW showed the strongest negative correlation with the global composite (ρ=−0.44, p<0.001); correlations for Ch1-3 GMD (ρ=0.38, p<0.001) and Ch1-3 FW (ρ=−0.39, p<0.001) paralleled those for MoCA.

**Figure 2.**
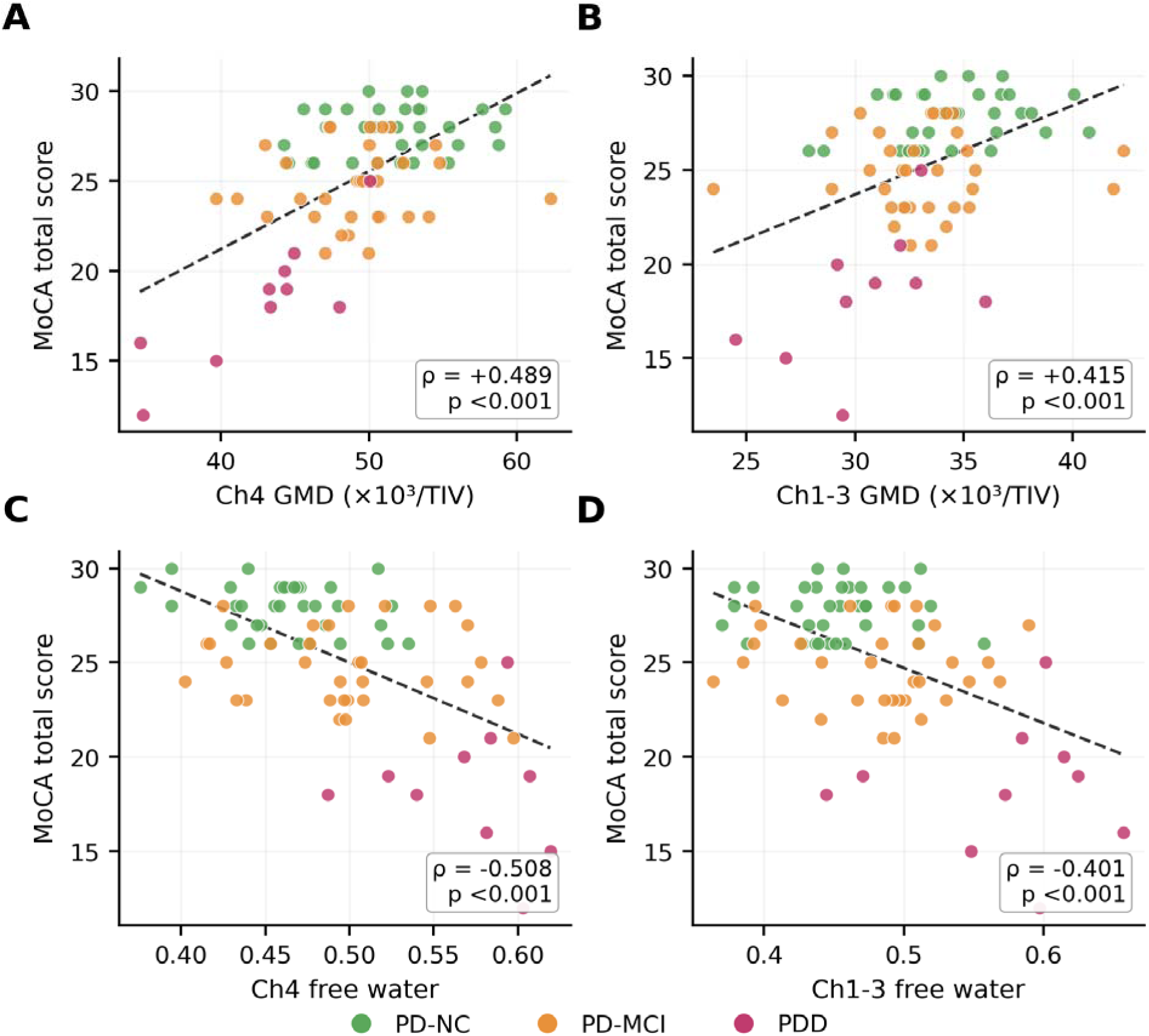
Scatter plots of cholinergic measures versus MoCA total score. A: Ch4 GMD; B: Ch1-3 GMD; C Ch4 free water; D: Ch1-3 free water. Dashed lines represent linear fit. Spearman ρ and p values shown.

Cognitive domain correlations of Ch4/Ch1-3 GMD and FW are summarized in Figure 3 (heatmap). Ch4 GMD and Ch4 FW showed the strongest and most consistent associations across cognitive domains, with the largest correlations observed for executive function (Ch4 GMD: ρ = +0.56, p < 0.001; Ch4 FW: ρ = −0.45, p < 0.001) and memory (ρ = +0.51 and −0.42, respectively; both p < 0.001). The visuospatial domain (represented by JLO z-score) showed weak, non-significant associations with all BF metrics (|ρ| ≤ 0.18). All significance markers in Figure 3 are uncorrected; all reported associations with p < 0.05 are also significant after Benjamini-Hochberg false-discovery-rate correction within row, except those for attention (for the GMD metrics) and visuospatial function (all metrics).

**Figure 3.**
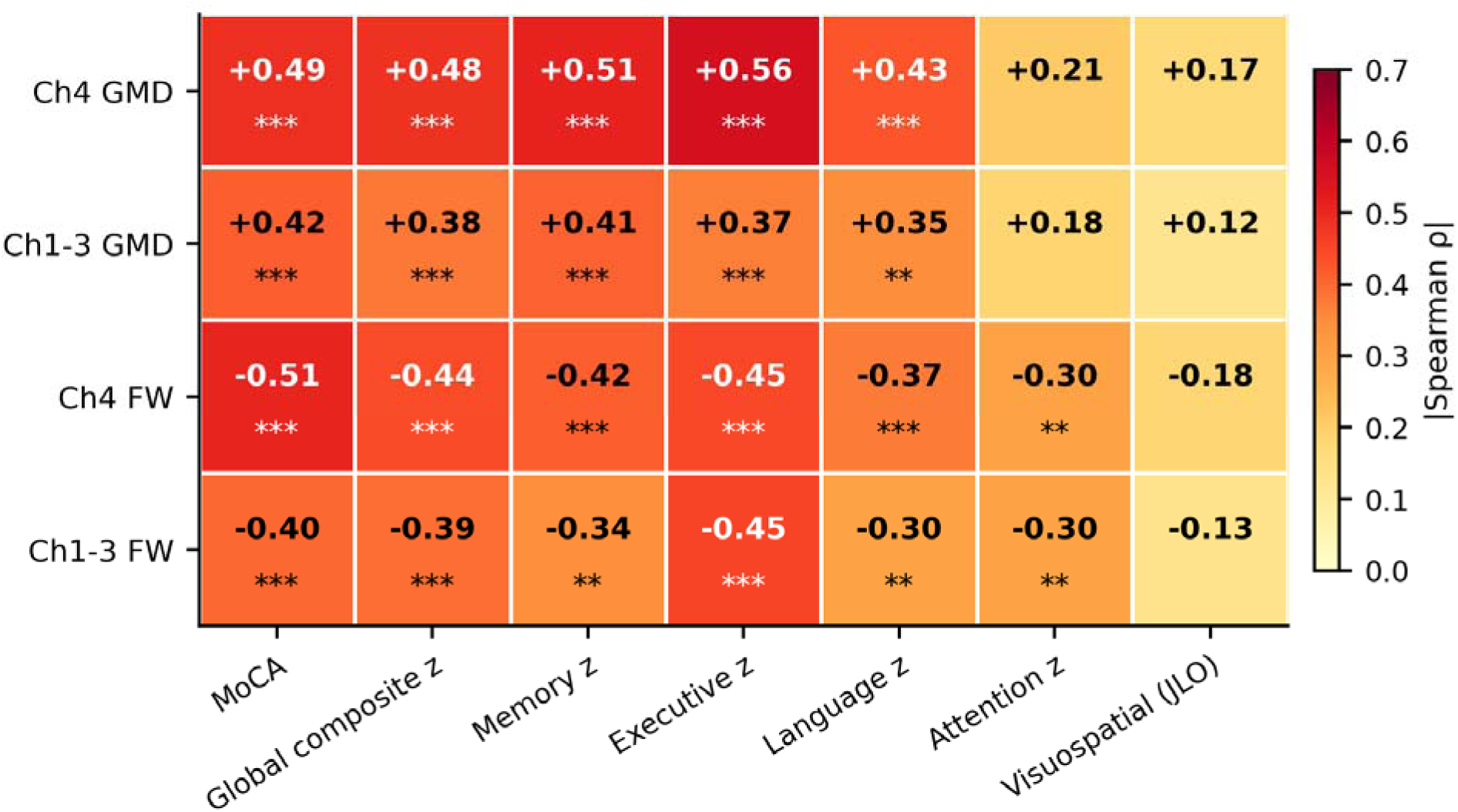
Heatmap of Spearman rank correlations between basal forebrain imaging metrics and cognitive measures in Parkinson’s disease (PD-only complete multimodal cohort, N = 78). Rows: four BF metrics. Columns: Montreal Cognitive Assessment (MoCA), global cognitive composite z-score, and five individual cognitive domain z-scores (memory, executive, language, attention, visuospatial (n=76)). Color intensity encodes |ρ|; signed ρ is printed in each cell. Significance markers reflect uncorrected p-values: *p < 0.05, **p < 0.01, ***p < 0.001. All effects with p < 0.05 also survive Benjamini-Hochberg false-discovery-rate correction within row except in the Attention (Ch4 GMD, Ch1-3 GMD rows) and Visuospatial (all rows) columns.

### Independent-predictor analyses

Using the tiered multivariable regression strategy described in the Methods, we examined which basal forebrain measures were independent predictors of cognition as measured by MoCA total score, the global composite z-score, and each of the five cognitive domain z-scores. The complete tier-by-tier estimates are reported in Supplementary Table S1; the main findings are summarized here.

Within the Ch4 nucleus model (Tier 2a), Ch4 GMD was the most predictive of MoCA score, the global composite z-score, and the memory, executive, language, and visuospatial domain z-scores. Ch4 FW added independent variance over Ch4 GMD for MoCA (β = −1.29 per SD, p = 0.002) and the attention domain (β = −0.25 per SD, p = 0.038) . Within the Ch1-3 nucleus model (Tier 2b), Ch1-3 FW was the most predictive for MoCA score, the global composite z-score, and the memory, executive, and language z-scores; Ch1-3 GMD was not a significant predictor of any cognitive outcome when included with Ch1-3 FW.

In the between-nucleus structural model (Tier 3a, Ch4 GMD + Ch1-3 GMD), Ch4 GMD was consistently more predictive than Ch1-3 GMD. In the between-nucleus microstructural model (Tier 3b, Ch4 FW + Ch1-3 FW), Ch4 FW retained the most consistent association across adjusted models, being the dominant microstructural predictor for MoCA, the global composite z-score, and language, with Ch1-3 FW comparable only for executive function.

In the full 4-metric model (Tier 4), Ch4 GMD remained the leading positive predictor for MoCA and the global composite, memory, executive, language, and visuospatial z-scores, and Ch4 FW remained an independent predictor for MoCA (β = −1.46 per SD, p = 0.005); Ch1-3 GMD emerged as an independent negative predictor for executive function (β = −0.50 per SD, p = 0.017) and for the global composite z-score (β = −0.31 per SD, p = 0.045), reflecting suppression by the more predictive Ch4 GMD term.

Taken together, these analyses identify Ch4 GMD as the most predictive of cognition in PD, with Ch4 FW as the greater microstructural predictor. The two-measure combination motivated by this pattern (Ch4 GMD + Ch4 FW, combining macrostructure and microstructure within the nucleus basalis of Meynert) is evaluated alongside other combinations in the discrimination analyses below.

### ROC analyses for cognitive classification

ROC analyses evaluated discrimination across two comparisons (Table 3, Figure 4). For classifying PDD versus non-demented PD patients (Panel A), the combined model showed high apparent discrimination, although estimates are unstable given only 10 PDD cases (AUC = 0.934 [0.858, 0.997]). For discriminating PD-CI (PDD + PD-MCI) from PD-NC (Panel B), the combined model achieved an AUC of 0.788 (95% CI [0.687, 0.886]), with Ch4 FW showing the highest single-feature AUC (0.765 [0.643, 0.865]). Across both classification tasks, the within-Ch4 combination Ch4 GMD + Ch4 FW was the best-performing two-feature model (PDD versus non-demented PD apparent AUC=0.934; PD-CI versus PD-NC apparent AUC=0.788).

**Figure 4.**
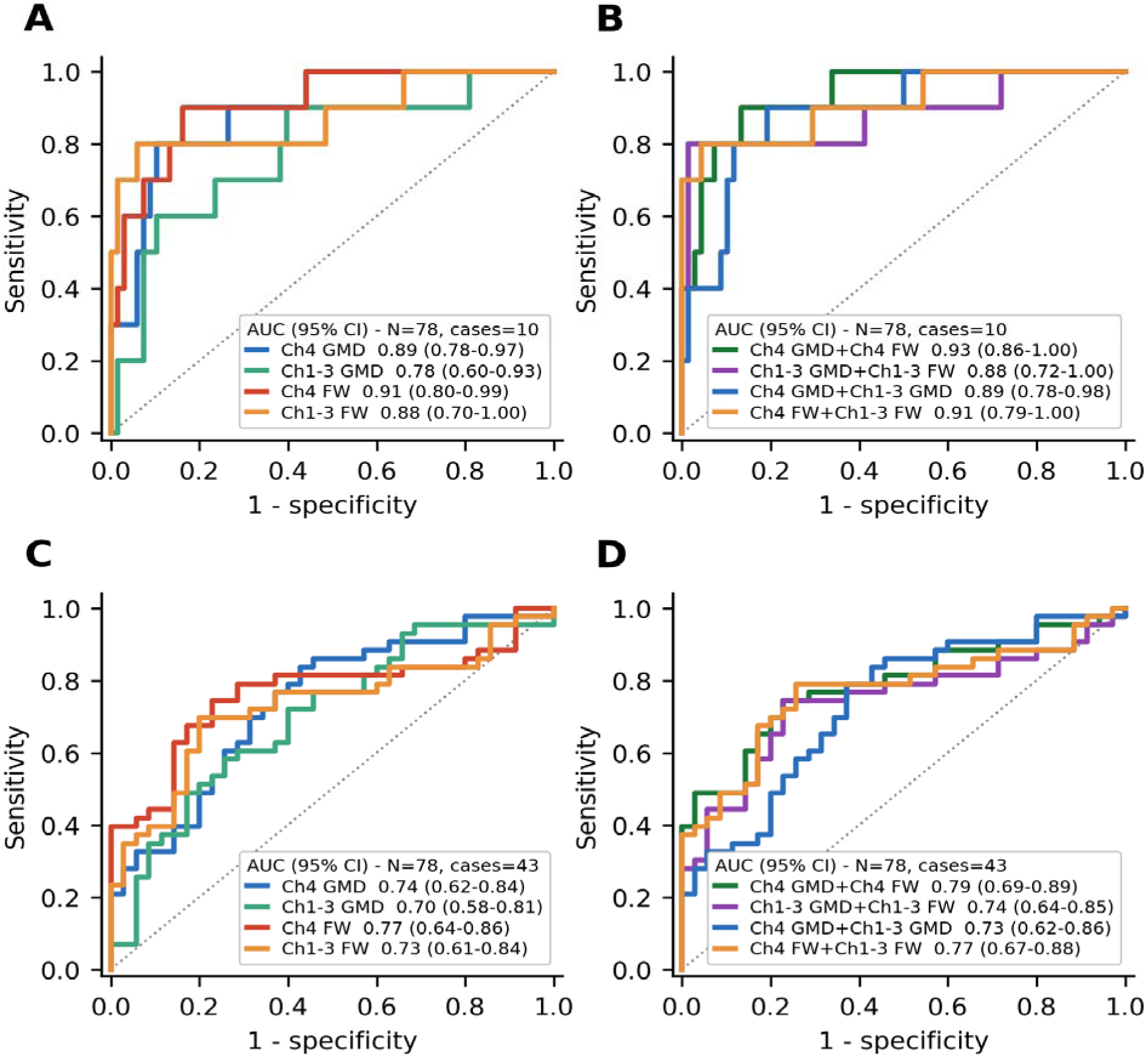
Receiver-operating-characteristic curves for binary classification of Parkinson’s disease cognitive stages by basal-forebrain imaging metrics. Top row: PDD versus non-demented PD (PD-NC + PD-MCI). Bottom row: PD with cognitive impairment (PD-MCI + PDD) versus PD with normal cognition. Left column (panels A and C): four individual BF metrics. Right column (panels B and D): four combined logistic-regression models — within-nucleus Ch4 (Ch4 GMD + Ch4 FW); within-nucleus Ch1-3 (Ch1-3 GMD + Ch1-3 FW); both GMD (Ch4 GMD + Ch1-3 GMD); and both FW (Ch4 FW + Ch1-3 FW). AUC values with 95% bootstrap confidence intervals shown in each panel. Sample: 78 PD patients with complete imaging data.

Bootstrap optimism correction (Harrell’s procedure, 1,000 resamples) produced negligible reductions in AUC: the combined Ch4 GMD + Ch4 FW model declined from 0.934 to 0.925 (Δ = 0.009) for PDD vs non-demented PD and from 0.788 to 0.773 (Δ = 0.015) for PD-CI vs PD-NC. The small magnitude of the corrections suggests limited apparent overfitting, although external validation remains essential.

## Discussion

This study provides a multimodal MRI assessment of cholinergic basal forebrain degeneration across cognitive stages in PD, integrating structural gray matter density with free-water metrics for both Ch4 and Ch1-3 nuclei. Our findings demonstrate that (1) both structural and diffusion measures of Ch4 and Ch1-3 integrity show stepwise changes across cognitive stages, (2) free water in Ch4 provides complementary discriminative information compared with structural volumetry alone, and (3) combining structural and diffusion markers showed high apparent discrimination for PDD (AUC = 0.93), requiring external validation.

The stepwise decline in Ch4 GMD across cognitive stages is consistent with the well-established role of Ch4 atrophy in PD-related cognitive decline (Barrett et al., 2019; Ray et al., 2018; Schulz et al., 2018). Our TIV-normalized Ch4 GMD values demonstrated a clear gradient (HC > PD-NC > PD-MCI > PDD), with PDD showing approximately 16% lower Ch4 GMD compared with PD-NC. These findings align with prior volumetric studies using similar probabilistic atlas-based approaches (Ray et al., 2018; Schulz et al., 2018). Notably, Ch1-3 also showed significant volume loss in PDD, supporting the notion that cognitive decline in PD involves broader cholinergic network degeneration beyond Ch4 (Zaborszky et al., 2008).

Within the PD spectrum, Ch4 FW was elevated in PDD compared with PD-NC (median 0.583 vs 0.467; 25% increase), with PD-MCI intermediate. Ch1-3 FW similarly showed a stepwise within-PD elevation, with PD-MCI exceeding PD-NC by 8% and PDD exceeding PD-NC by 30% (Figure 1). Our FW results extend a small but growing body of work applying free-water diffusion imaging to the cholinergic BF in PD. Ray and colleagues reported elevated FW in the cholinergic BF and pedunculopontine nucleus in PD, with BF FW correlating with cognitive performance (Ray et al., 2023). Zhang et al. similarly demonstrated increased NBM free water in both isolated REM sleep behavior disorder and established PD, supporting NBM FW as an early marker preceding overt atrophy (Zhang et al., 2024). Wu et al. linked cholinergic BF free water to postural instability/gait difficulty and attentional impairment in PD (Wu et al., 2024), and Xu et al. recently reported FW associations with motor subtypes (Xu et al., 2025). Beyond the BF, FW elevation has been demonstrated in the substantia nigra (Ofori et al., 2015) and as a progression marker in PD (Burciu et al., 2017). Our data converge with and extend these reports in three respects: (i) we observed a stepwise (HC → PD-NC → PD-MCI → PDD) FW gradient, where prior work has mostly contrasted PD vs HC or single PD groups; (ii) the magnitude of the Ch4 FW elevation in PDD vs PD-NC (∼25%) is consistent with the effect sizes reported by Ray et al.(Ray et al., 2023) and Zhang et al.(Zhang et al., 2024); and (iii) Ch1-3 FW showed a comparable, slightly larger PDD-vs-PD-NC change (∼30%), supporting cholinergic-network-wide microstructural disruption rather than NBM-only pathology — a pattern consistent with neuropathological evidence of Ch1-3 involvement in PDD (Liu et al., 2015).

Cognitive correlations followed a coherent neurobiological pattern. MoCA and the global composite z-score tracked together, as expected given their overlapping content. Executive, memory, and language domains each correlated with both structural (Ch4/Ch1-3 GMD) and microstructural (Ch4/Ch1-3 FW) BF measures, consistent with the dependence of these domains on widely distributed cortical projections of NBM origin (Bohnen & Albin, 2011; Pasquini et al., 2021). The language composite includes category fluency, which has a substantial executive/frontal-systems component (Henry & Crawford, 2004a, 2004b), and likely contributes to the parallel pattern observed across executive and language domains. Of note, attention correlated with FW metrics but not consistently with GMD; one interpretation is that attentional networks are sensitive to early microstructural disruption (neuroinflammation, extracellular-water expansion) before overt volumetric loss accrues, paralleling cholinergic-PET findings of attention–acetylcholine coupling in PD (Bohnen et al., 2006; Bohnen & Albin, 2011; Pasquini et al., 2021).

Beyond group-level differences, multivariable regression analyses identified Ch4 GMD as the most predictive independent structural correlate of cognition for MoCA, the global composite z-score, and most cognitive domains, and Ch4 FW as the most predictive independent microstructural feature (Supplementary File). GMD and FW represent distinct, partially independent aspects of BF degeneration. GMD is a macrostructural volumetric measure sensitive to neuronal loss and atrophy (Barrett et al., 2019; Ray et al., 2018; Schulz et al., 2018), whereas FW estimates the extracellular water compartment and is sensitive to tissue rarefaction, neuroinflammation, and disruption of cell-membrane density that may precede overt volume loss (Haddad et al., 2025; Ofori et al., 2015; Pasternak et al., 2009; Ray et al., 2023; Zhang et al., 2024). Their partial independence is supported by prior PD literature showing that nigral free water rises before substantial volumetric change (Archer et al., 2019; Burciu et al., 2016; Ofori et al., 2015) and by our own finding that, in head-to-head 4-metric models, Ch4 GMD and Ch4 FW both retained independent associations with MoCA (Tier 4). The Ch4 GMD + Ch4 FW pairing thus captures complementary macrostructural and microstructural degeneration within the nucleus basalis of Meynert, and is the parsimonious operationalization of multimodal BF integrity in our data. The persistence of Ch4 GMD as the leading positive predictor in the full 4-metric model is consistent with neuropathological evidence placing the nucleus basalis of Meynert (Ch4) as the principal cholinergic source for cortical projections relevant to cognition in PD (Pasquini et al., 2021; Ray et al., 2018; Schulz et al., 2018).

Domain-specific multivariable findings reinforced the leading role of Ch4 GMD. Across MoCA and the memory, executive, language, and visuospatial domains, Ch4 GMD was the strongest independent positive predictor, whereas Ch4 FW retained an independent association specifically with global MoCA performance. Ch4 FW emerged as the leading microstructural predictor, consistent with the nucleus basalis of Meynert serving as the principal cholinergic source for cortical projections relevant to cognition (Bohnen & Albin, 2011; Pasquini et al., 2021). The visuospatial domain, represented by JLO alone (see Methods), showed effects consistent with the other domains but with broader confidence intervals.

The apparent negative β-coefficient for Ch1-3 GMD in the 4-metric model should not be interpreted as larger Ch1-3 volume conferring worse cognition. Ch4 and Ch1-3 GMD are highly collinear because BF nuclei degenerate concomitantly in PD (Grothe et al., 2021; Ray et al., 2018; Schulz et al., 2018); when both are entered jointly, the variance shared with cognition is absorbed by the stronger predictor (Ch4 GMD), and the partial Ch1-3 GMD coefficient reflects the residual between-nucleus contrast (Ch1-3 being relatively spared given the level of Ch4 atrophy). This is a classical multicollinear-suppression pattern (MacKinnon et al., 2000). Univariate associations of Ch1-3 GMD with cognition remain positive (Table 2), as expected biologically.

**Table 2.** Spearman correlations between cholinergic basal forebrain measures and MoCA in Parkinson’s disease.

| Measure | Spearman $\rho$ | p-value |
| --- | --- | --- |
| Ch4 GMD | +0.489 | <0.001 |
| Ch1-3 GMD | +0.415 | <0.001 |
| Ch4 FW | –0.508 | <0.001 |
| Ch1-3 FW | –0.401 | <0.001 |

All Spearman rank correlations were computed within the N = 78 Parkinson's disease participants with complete multimodal basal forebrain imaging (Ch4 GMD, Ch1-3 GMD, Ch4 FW, and Ch1-3 FW all available). Abbreviations: FW, free water fraction; GMD, gray matter density; MoCA, Montreal Cognitive Assessment.

**Table 3.** Receiver-operating-characteristic analysis for classifying Parkinson’s disease cognitive stages from basal forebrain imaging metrics.

| Feature | AUC (95% CI) | Sensitivity | Specificity | N |
| --- | --- | --- | --- | --- |
| **PDD vs non-demented PD** |  |  |  |  |
| Ch4 GMD | 0.891 (0.778–0.973) | 0.800 | 0.897 | 78 |
| Ch1-3 GMD | 0.782 (0.597–0.926) | 0.900 | 0.603 | 78 |
| Ch4 FW | 0.912 (0.798–0.986) | 0.900 | 0.838 | 78 |
| Ch1-3 FW | 0.876 (0.700–0.999) | 0.800 | 0.941 | 78 |
| Combined (Ch4 GMD + Ch4 FW) | 0.934 (0.858–0.997) | 0.900 | 0.868 | 78 |
| Combined (Ch1-3 GMD + Ch1-3 FW) | 0.881 (0.716–1.000) | 0.800 | 0.985 | 78 |
| Combined (Ch4 GMD + Ch1-3 GMD) | 0.887 (0.782–0.980) | 0.900 | 0.809 | 78 |
| Combined (Ch4 FW + Ch1-3 FW) | 0.912 (0.794–1.000) | 0.800 | 0.956 | 78 |
| **PD-CI vs PD-NC** |  |  |  |  |
| Ch4 GMD | 0.738 (0.617–0.845) | 0.837 | 0.571 | 78 |
| Ch1-3 GMD | 0.700 (0.583–0.807) | 0.581 | 0.743 | 78 |
| Ch4 FW | 0.765 (0.643–0.865) | 0.744 | 0.771 | 78 |
| Ch1-3 FW | 0.730 (0.615–0.842) | 0.698 | 0.800 | 78 |
| Combined (Ch4 GMD + Ch4 FW) | 0.788 (0.687–0.886) | 0.744 | 0.771 | 78 |
| Combined (Ch1-3 GMD + Ch1-3 FW) | 0.741 (0.641–0.852) | 0.744 | 0.771 | 78 |
| Combined (Ch4 GMD + Ch1-3 GMD) | 0.734 (0.623–0.856) | 0.791 | 0.629 | 78 |
| Combined (Ch4 FW + Ch1-3 FW) | 0.770 (0.665–0.875) | 0.791 | 0.743 | 78 |

AUC 95% confidence intervals were estimated using 2,000 bootstrap resamples; optimism correction used Harrell's bootstrap procedure with 1,000 resamples. Bootstrap optimism-corrected AUCs for the combined models: Ch4 GMD + Ch4 FW = 0.925 (PDD vs non-demented PD) and 0.773 (PD-CI vs PD-NC); Ch1-3 GMD + Ch1-3 FW = 0.865 and 0.726; Ch4 GMD + Ch1-3 GMD = 0.875 and 0.716; Ch4 FW + Ch1-3 FW = 0.904 and 0.758. Sensitivity and specificity reported at the optimal Youden index threshold. Combined models entered the listed structural and diffusion features simultaneously as predictors in logistic regression. Abbreviations: AUC, area under the receiver-operating-characteristic curve; FW, free water fraction; GMD, gray matter density; PD-CI, Parkinson's disease with cognitive impairment (PD-MCI + PDD); PD-NC, Parkinson's disease with normal cognition; PDD, Parkinson's disease with dementia.

Sensitivity analyses restricted to non-demented PD (PD-NC + PD-MCI; Supplementary File) confirmed that Ch4 GMD, Ch1-3 GMD, and Ch4 FW remained significantly correlated with MoCA and the global composite z-score even with PDD excluded (Ch1-3 FW attenuated to borderline significance), indicating that the BF–cognition relationship is present along the PD cognitive spectrum and is not a PDD-specific artifact. As expected for a continuous biological gradient compressed by removing its tail, effect sizes attenuated in the non-demented subset, reaffirming that statistical detection of BF imaging effects is most powered when the full cognitive spectrum (including PDD) is sampled.

Our ROC analyses provide preliminary evidence that combining structural and diffusion markers could improve discrimination. While Ch4 GMD alone showed promising exploratory discrimination of PDD (AUC = 0.891), Ch4 FW performed even better as a single feature (AUC = 0.912). Their combination (AUC = 0.934, sensitivity 90%, specificity 87%) suggests that structural and diffusion measures capture partially non-overlapping aspects of Ch4 degeneration. Notably, when all four cholinergic basal forebrain metrics were entered simultaneously, Ch4 GMD and Ch4 FW emerged as the only independent predictors of MoCA, while Ch1-3 measures did not contribute additional predictive information, suggesting that Ch4 drives the cognitive association in this multimodal framework. For the broader distinction of cognitively impaired PD (PD-MCI + PDD) from PD-NC, the combined model achieved an AUC of 0.788, reflecting the subtler neurobiological boundary between intact and mildly impaired cognition in PD.

Several limitations should be acknowledged. First, the cross-sectional design precludes assessment of longitudinal trajectories, and longitudinal data will be required to determine whether free-water changes precede atrophy or predict conversion from PD-MCI to PDD. Second, the PDD subgroup was modest (N = 10); although bootstrap optimism correction produced only a minimal reduction in the AUCs (≤0.018 across all combined models), suggesting the discrimination is not predominantly driven by overfitting; nevertheless, classification estimates require external validation in larger independent samples. Third, disease duration, levodopa-equivalent daily dose, and cholinesterase-inhibitor exposure were not available for all participants. Fourth, the visuospatial domain was represented by the Judgment of Line Orientation z-score alone, as the cohort harmonization plan did not allow a composite visuospatial z-score (see Methods). Fifth, free-water fraction was estimated from b = 0 and b = 1000 data using the bi-tensor model; single-shell free-water estimation is more model-constrained than multi-shell approaches and should be interpreted with appropriate caution.

In conclusion, in this cross-sectional single-center cohort, lower Ch4 GMD and higher basal forebrain FW were associated with worse cognition across the PD cognitive spectrum. Tiered analyses demonstrated that structural (Ch4 GMD) and microstructural (Ch4 FW) measures of the nucleus basalis of Meynert carry complementary, non-redundant information about cholinergic system integrity relevant to cognition in PD. Exploratory classification analyses suggested that combining structural and free-water metrics may improve discrimination of PDD, but the small PDD subgroup and absence of external validation require cautious interpretation. Future longitudinal studies are needed to determine whether free-water changes precede atrophy and predict conversion from PD-MCI to PDD, and whether free water in Ch4 (reflecting extracellular-water expansion from neurodegeneration, neuroinflammation, or atrophy-associated tissue rarefaction) adds independent prognostic information beyond structural volumetry.

## Data Availability

All data produced in the present study are available upon reasonable request to the authors (Ahmed Negida and Matthew Barrett).

## Declarations

### Ethics

All study procedures were approved by the Virginia Commonwealth University Institutional Review Board and all participants provided written informed consent or consent was obtained from a legally authorized representative when applicable.

## Acknowledgements

We thank the participants and their families for their contributions to this research, and the staff of the Virginia Commonwealth University Parkinson and Movement Disorder Center for their assistance with data collection.

## Author contributions

A.N. conceived and designed the study, performed the statistical analyses, and drafted the manuscript. R.H., A.Z., K.W.-C., C.M.-P., B.D.B., E.O., and M.J.B. contributed to data acquisition, interpretation of results, and critical revision of the manuscript. M.J.B. supervised the study. All authors approved the final version of the manuscript.

## Data availability

De-identified imaging and cognitive data supporting the findings of this study are available from the corresponding author upon reasonable request and approval by the Virginia Commonwealth University Institutional Review Board, subject to a data sharing agreement.

