## Supplementary material for "Multimodal MRI Characterization of Nucleus Basalis of Meynert Degeneration: Structural Atrophy and Free-water Diffusion in Parkinson’s Disease Cognitive Impairment": Supplementary File.docx

### **Additional Results 1: Sensitivity analyses in non-demented Parkinson’s disease**

To verify that the basal forebrain–cognition relationships were not driven exclusively by Parkinson’s disease dementia (PDD), the correlational analyses were repeated in the Parkinson’s disease with normal cognition (PD-NC) and Parkinson’s disease with mild cognitive impairment (PD-MCI) participants only (non-demented PD, N = 68).

Spearman rank correlations with the Montreal Cognitive Assessment (MoCA) remained significant for Ch4 GMD (ρ = +0.30, *p* = 0.012), Ch1-3 GMD (ρ = +0.30, *p* = 0.013), and Ch4 FW (ρ = −0.34, p = 0.004); Ch1-3 FW attenuated to borderline significance (ρ = −0.24, p = 0.053). A similar pattern was observed for the global cognitive composite z-score: Ch4 GMD (ρ = +0.31, *p* = 0.010), Ch1-3 GMD (ρ = +0.27, *p* = 0.028), Ch4 FW (ρ = −0.27, *p* = 0.025), and Ch1-3 FW (ρ = −0.24, *p* = 0.050).

These results indicate that the basal forebrain–cognition relationship is present along the Parkinson’s disease cognitive spectrum and is not a PDD-specific artifact. Ch4 FW remained the most consistent microstructural correlate of cognition in the non-demented subset, whereas Ch1-3 FW attenuated to borderline significance. As expected for a continuous biological gradient compressed by removing its severe tail, effect sizes were attenuated relative to the full cohort, reaffirming that statistical detection of basal forebrain imaging effects is most powered when the full cognitive spectrum (including PDD) is sampled.

**Supplementary Table S1.** Tiered multivariable linear regression of cholinergic basal forebrain imaging metrics on cognitive outcomes in Parkinson’s disease.

| **Tier (model)** | **Outcome** | **Predictor** | **β (per SD)** | **p-value** | **Adj. R²** |
| --- | --- | --- | --- | --- | --- |
| **Tier 1 (univariate): Ch4 GMD** | MoCA total | Ch4 GMD | +2.03 | <0.001 | 0.365 |
|  | Global composite z | Ch4 GMD | +0.50 | <0.001 | 0.270 |
|  | Memory z | Ch4 GMD | +0.66 | <0.001 | 0.287 |
|  | Executive z | Ch4 GMD | +0.68 | <0.001 | 0.262 |
|  | Language z | Ch4 GMD | +0.46 | <0.001 | 0.210 |
|  | Attention z | Ch4 GMD | +0.20 | 0.052 | 0.056 |
|  | Visuospatial z (JLO) | Ch4 GMD | +0.47 | 0.012 | 0.089 |
| **Tier 1 (univariate): Ch1-3 GMD** | MoCA total | Ch1-3 GMD | +1.34 | 0.001 | 0.201 |
|  | Global composite z | Ch1-3 GMD | +0.30 | 0.009 | 0.097 |
|  | Memory z | Ch1-3 GMD | +0.42 | 0.005 | 0.136 |
|  | Executive z | Ch1-3 GMD | +0.38 | 0.015 | 0.079 |
|  | Language z | Ch1-3 GMD | +0.30 | 0.006 | 0.072 |
|  | Attention z | Ch1-3 GMD | +0.09 | 0.399 | 0.015 |
|  | Visuospatial z (JLO) | Ch1-3 GMD | +0.25 | 0.199 | 0.026 |
| **Tier 1 (univariate): Ch4 FW** | MoCA total | Ch4 FW | -2.00 | <0.001 | 0.345 |
|  | Global composite z | Ch4 FW | -0.46 | <0.001 | 0.218 |
|  | Memory z | Ch4 FW | -0.57 | <0.001 | 0.218 |
|  | Executive z | Ch4 FW | -0.61 | <0.001 | 0.201 |
|  | Language z | Ch4 FW | -0.43 | <0.001 | 0.173 |
|  | Attention z | Ch4 FW | -0.29 | 0.006 | 0.105 |
|  | Visuospatial z (JLO) | Ch4 FW | -0.35 | 0.066 | 0.050 |
| **Tier 1 (univariate): Ch1-3 FW** | MoCA total | Ch1-3 FW | -1.65 | <0.001 | 0.268 |
|  | Global composite z | Ch1-3 FW | -0.42 | <0.001 | 0.189 |
|  | Memory z | Ch1-3 FW | -0.52 | <0.001 | 0.193 |
|  | Executive z | Ch1-3 FW | -0.62 | <0.001 | 0.217 |
|  | Language z | Ch1-3 FW | -0.36 | <0.001 | 0.118 |
|  | Attention z | Ch1-3 FW | -0.26 | 0.010 | 0.092 |
|  | Visuospatial z (JLO) | Ch1-3 FW | -0.32 | 0.087 | 0.044 |
| **Tier 2a (within-Ch4): Ch4 GMD + Ch4 FW** | MoCA total | Ch4 GMD | +1.41 | <0.001 | 0.439 |
|  |  | Ch4 FW | -1.29 | 0.002 | 0.439 |
|  | Global composite z | Ch4 GMD | +0.37 | 0.001 | 0.315 |
|  |  | Ch4 FW | -0.27 | 0.018 | 0.315 |
|  | Memory z | Ch4 GMD | +0.51 | <0.001 | 0.319 |
|  |  | Ch4 FW | -0.32 | 0.039 | 0.319 |
|  | Executive z | Ch4 GMD | +0.51 | 0.001 | 0.302 |
|  |  | Ch4 FW | -0.35 | 0.026 | 0.302 |
|  | Language z | Ch4 GMD | +0.33 | 0.003 | 0.256 |
|  |  | Ch4 FW | -0.26 | 0.022 | 0.256 |
|  | Attention z | Ch4 GMD | +0.08 | 0.490 | 0.098 |
|  |  | Ch4 FW | -0.25 | 0.038 | 0.098 |
|  | Visuospatial z (JLO) | Ch4 GMD | +0.40 | 0.060 | 0.084 |
|  |  | Ch4 FW | -0.16 | 0.449 | 0.084 |
| **Tier 2b (within-Ch1-3): Ch1-3 GMD + Ch1-3 FW** | MoCA total | Ch1-3 GMD | +0.56 | 0.242 | 0.272 |
|  |  | Ch1-3 FW | -1.33 | 0.006 | 0.272 |
|  | Global composite z | Ch1-3 GMD | +0.08 | 0.552 | 0.182 |
|  |  | Ch1-3 FW | -0.37 | 0.005 | 0.182 |
|  | Memory z | Ch1-3 GMD | +0.18 | 0.313 | 0.193 |
|  |  | Ch1-3 FW | -0.42 | 0.016 | 0.193 |
|  | Executive z | Ch1-3 GMD | +0.02 | 0.896 | 0.206 |
|  |  | Ch1-3 FW | -0.61 | <0.001 | 0.206 |
|  | Language z | Ch1-3 GMD | +0.14 | 0.284 | 0.120 |
|  |  | Ch1-3 FW | -0.28 | 0.030 | 0.120 |
|  | Attention z | Ch1-3 GMD | -0.10 | 0.431 | 0.088 |
|  |  | Ch1-3 FW | -0.32 | 0.011 | 0.088 |
|  | Visuospatial z (JLO) | Ch1-3 GMD | +0.09 | 0.711 | 0.032 |
|  |  | Ch1-3 FW | -0.27 | 0.237 | 0.032 |
| **Tier 3a (between-nucleus): Ch4 GMD + Ch1-3 GMD** | MoCA total | Ch4 GMD | +2.45 | <0.001 | 0.364 |
|  |  | Ch1-3 GMD | -0.55 | 0.327 | 0.364 |
|  | Global composite z | Ch4 GMD | +0.66 | <0.001 | 0.280 |
|  |  | Ch1-3 GMD | -0.22 | 0.156 | 0.280 |
|  | Memory z | Ch4 GMD | +0.82 | <0.001 | 0.288 |
|  |  | Ch1-3 GMD | -0.21 | 0.307 | 0.288 |
|  | Executive z | Ch4 GMD | +0.94 | <0.001 | 0.281 |
|  |  | Ch1-3 GMD | -0.35 | 0.093 | 0.281 |
|  | Language z | Ch4 GMD | +0.55 | <0.001 | 0.206 |
|  |  | Ch1-3 GMD | -0.12 | 0.423 | 0.206 |
|  | Attention z | Ch4 GMD | +0.32 | 0.046 | 0.055 |
|  |  | Ch1-3 GMD | -0.16 | 0.329 | 0.055 |
|  | Visuospatial z (JLO) | Ch4 GMD | +0.66 | 0.019 | 0.088 |
|  |  | Ch1-3 GMD | -0.26 | 0.358 | 0.088 |
| **Tier 3b (between-nucleus): Ch4 FW + Ch1-3 FW** | MoCA total | Ch4 FW | -1.65 | 0.003 | 0.343 |
|  |  | Ch1-3 FW | -0.48 | 0.366 | 0.343 |
|  | Global composite z | Ch4 FW | -0.32 | 0.039 | 0.226 |
|  |  | Ch1-3 FW | -0.19 | 0.195 | 0.226 |
|  | Memory z | Ch4 FW | -0.40 | 0.053 | 0.223 |
|  |  | Ch1-3 FW | -0.24 | 0.232 | 0.223 |
|  | Executive z | Ch4 FW | -0.31 | 0.124 | 0.232 |
|  |  | Ch1-3 FW | -0.39 | 0.051 | 0.232 |
|  | Language z | Ch4 FW | -0.35 | 0.023 | 0.168 |
|  |  | Ch1-3 FW | -0.11 | 0.443 | 0.168 |
|  | Attention z | Ch4 FW | -0.19 | 0.189 | 0.102 |
|  |  | Ch1-3 FW | -0.12 | 0.388 | 0.102 |
|  | Visuospatial z (JLO) | Ch4 FW | -0.24 | 0.390 | 0.040 |
|  |  | Ch1-3 FW | -0.15 | 0.586 | 0.040 |
| **Tier 4 (full 4-metric)** | MoCA total | Ch4 GMD | +2.04 | <0.001 | 0.446 |
|  |  | Ch1-3 GMD | -0.90 | 0.101 | 0.446 |
|  |  | Ch4 FW | -1.46 | 0.005 | 0.446 |
|  |  | Ch1-3 FW | +0.08 | 0.882 | 0.446 |
|  | Global composite z | Ch4 GMD | +0.56 | <0.001 | 0.335 |
|  |  | Ch1-3 GMD | -0.31 | 0.045 | 0.335 |
|  |  | Ch4 FW | -0.27 | 0.055 | 0.335 |
|  |  | Ch1-3 FW | -0.07 | 0.648 | 0.335 |
|  | Memory z | Ch4 GMD | +0.70 | 0.001 | 0.322 |
|  |  | Ch1-3 GMD | -0.31 | 0.139 | 0.322 |
|  |  | Ch4 FW | -0.33 | 0.085 | 0.322 |
|  |  | Ch1-3 FW | -0.05 | 0.819 | 0.322 |
|  | Executive z | Ch4 GMD | +0.77 | <0.001 | 0.345 |
|  |  | Ch1-3 GMD | -0.50 | 0.017 | 0.345 |
|  |  | Ch4 FW | -0.27 | 0.161 | 0.345 |
|  |  | Ch1-3 FW | -0.26 | 0.204 | 0.345 |
|  | Language z | Ch4 GMD | +0.47 | 0.003 | 0.253 |
|  |  | Ch1-3 GMD | -0.19 | 0.214 | 0.253 |
|  |  | Ch4 FW | -0.30 | 0.038 | 0.253 |
|  |  | Ch1-3 FW | +0.02 | 0.884 | 0.253 |
|  | Attention z | Ch4 GMD | +0.21 | 0.196 | 0.110 |
|  |  | Ch1-3 GMD | -0.25 | 0.120 | 0.110 |
|  |  | Ch4 FW | -0.20 | 0.182 | 0.110 |
|  |  | Ch1-3 FW | -0.14 | 0.366 | 0.110 |
|  | Visuospatial z (JLO) | Ch4 GMD | +0.61 | 0.041 | 0.073 |
|  |  | Ch1-3 GMD | -0.30 | 0.299 | 0.073 |
|  |  | Ch4 FW | -0.21 | 0.445 | 0.073 |
|  |  | Ch1-3 FW | +0.01 | 0.959 | 0.073 |

*All models were fit in the N = 78 Parkinson’s disease participants with complete multimodal basal forebrain imaging and adjusted for age, sex, and education; imaging predictors were z-standardized within the Parkinson’s disease sample, so β denotes the change in outcome per 1-SD increase in the predictor. Tier 1 entered each metric alone; Tier 2 paired gray matter density and free water within the same nucleus; Tier 3 paired the two structural (3a) or the two microstructural (3b) metrics across nuclei; Tier 4 entered all four metrics simultaneously (coefficients subject to collinearity-driven instability). Abbreviations: FW, free water fraction; GMD, gray matter density; SD, standard deviation; MoCA, Montreal Cognitive Assessment; JLO, Judgment of Line Orientation.*
